# Variant Creutzfeldt-Jakob Disease: The Challenge of Preventing a Rare but Potentially Devastating Exposure

**DOI:** 10.1101/2021.04.08.21255124

**Authors:** Ritambhara Pandey, Devesh Rai, Julie Giles, Maryrose Laguio-Vila, Emil Lesho

**Affiliations:** Rochester Regional Health, Rochester, NY, USA

## Abstract

Although rare, patients with variant Creutzfeldt-Jakob Disease (vCJD) in their differential diagnosis of progressive dementia and movement disorder could continue to present to hospitals for care. However, U.S.-based infection control guidelines do not fully address the possibility of vCJD. After near-misses involving increasing numbers of patients with clinical findings and epidemiologic risks compatible with vCJD, or exposures to chronic wasting disease, we sought to improve recognition and prevention of iatrogenic spread of these prion-related diseases.

## Introduction

Transmissible spongiform encephalopathies (TSEs) are progressively fatal neurodegenerative diseases caused by infectious, misshapen prion proteins (PrP^Sc^). In humans, they include variant and classic (or sporadic) Creutzfeldt-Jakob Disease (vCJD/sCJD), the latter being far more common in the U.S. Animal forms of prion disease include bovine spongiform encephalopathy (BSE) and chronic wasting disease in cervids and some rodents.

TSEs present substantial diagnostic, infection control, and patient notification challenges detailed in the discussion section below. Because prions resist usual standard disinfection and sterilization protocols, iatrogenic exposures can occur via contaminated instruments or equipment. Iatrogenic TSE can also be transmitted by percutaneous exposure to certain types of tissue (mostly neural for sCJD), while vCJD can also be transmitted by exposure to infected lymphoid tissue and possibly blood transfusions. This is why people from regions where BSE was prevalent are still precluded from donating blood.^1-3^

However, U.S.-based cleaning and disinfection and control guidelines and updates do not address the possibility of vCJD.^4, 5^ One of the reasons it is not included in general procedures is that it is a rare occurrence and a special consideration. vCJD involves separate procedures. That are not applied across all reprocessing procedures, and different types of tissue that are potentially infectious (not only neural).

Those guidelines do, however, recommend heightened awareness and policy modification as events dictate.^4, 5^ Additionally, experts have noted persistent risk of TSEs other than sCJD in unsuspected geographic locations and a need for heightened suspicion and surveillance.^6, 7^ Furthermore, surveys of lymphoid tissue where PrP^sc^ is found in cases of vCJD, suggest the prevalence of asymptomatic disease can be as high as 1 in 2,000 in persons born between 1941 and 1985.^8^

### Two Illustrative Incidents

Two incidents typified the challenges we faced in caring for these and similar patients and provided the impetus for this approach. The first incident involved a patient who presented with memory problems, apathy and depression leading to job loss. These symptoms were followed by confusion, impaired gait, occasional jerking movements and eventually dementia. The patient lived in England and France from 1985 to 1993 and ate canned dog food occasionally. The patient also required urgent pelvic and abdominal surgery immediately after admission. MRI showed restricted diffusion in the frontal, parietal, and occipital cortexes. 14-3-3, and Tau protein were negative. Since immune mediated encephalitis was also a leading consideration on the list of possible diagnoses, the patient underwent plasma exchange. 13 days later, the RTQuIC result became available and was positive. Contact tracing revealed that in the meantime, three other patients had undergone pheresis on the same machine. The Red Cross was contacted and recommended decommissioning the machine until further notice. The dilemma of patient notification was debated by the ethics, risk management, infection control team and the hospital leaders (Table 1).

**Table 1:** Diagnostic, Infection Control, and Notification Challenges Associated with Transmissible Spongiform Encephalopathies.

|  |
| --- |
| **Diagnostic Challenge** |
| Most clinicians are unfamiliar with TSEs due to the rarity of those diseases. |
| vCJD and CJD can be hard to distinguish clinically. |
| No gold standard laboratory test exists for differentiating types of TSEs. |
| Turn-around-time for RT-QiC can be up to 14 days or longer. |
| In some cases, definitive diagnosis requires brain biopsy or only becomes available after autopsy. |
| The National Prion Center typically only releases autopsy results to public health departments, furthering delay. |
| **Infection Control Challenges** |
| Prions resist usual disinfection and sterilization methods. |
| US-based disinfection and sterilization guidelines do not address possibility of vCJD contaminated equipment. |
| **Notification Challenges or Considerations** |
| Should potentially exposed patients be informed given the below considerations? |
| i. Potential negative impact of informing patients about a fatal, untreatable brain disease. |
| ii. No effective screening test is available for TSE. |
| iii. No post exposure prophylaxis is available. |
| iv. Contaminated instruments often become mixed during re-processing making identification and linkage to specific patients nearly impossible. |
| v. Autoclaving is generally superior to chemical reprocessing for prions. |
| vi. If equipment was re-processed more than twice, prions would be less likely to survive. |

**Table 2:** Infectivity of Specific Types of Tissue.

|  | **Classic/ Sporadic CJD** | **Variant CJD** |
| --- | --- | --- |
| **High Infectivity Tissue**<br>**NO Reprocessing Allowed\*** | Brain (including dura mater)<br>Spinal cord<br>Posterior eye<br>Pituitary tissue | Brain (including dura mater)<br>Spinal cord, spinal ganglia<br>Posterior eye<br>Pituitary tissue<br>Trigeminal ganglia |
| **Low Infectivity Tissue**<br>**NO Reprocessing Allowed\*** | Cerebrospinal fluid (CSF)<br>Liver<br>Lymph node<br>Kidney<br>Lung (Bronchoscope)<br>Spleen<br>Placenta<br>Olfactory epithelium | Adrenal gland<br>Blood<br>Tonsils (Laryngoscope)<br>Cerebrospinal fluid (CSF)<br>Intestine (Endoscope)<br>Bone marrow<br>Liver<br>Lymph node<br>Kidney<br>Lung (Bronchoscope)<br>Spleen<br>Skeletal muscle<br>Placenta<br>Olfactory epithelium<br>Peripheral Nerves<br>Rectum (Endoscope) |
| **Tissue with no detectable infectivity**<br>**OK to Reprocess\*** | Peripheral nerve<br>Intestine<br>Bone marrow<br>Whole blood, leukocytes, serum, plasma<br>Thyroid gland<br>Adrenal gland<br>Heart<br>Skeletal muscle<br>Adipose tissue<br>Gingiva<br>Prostate and testis<br>Tears, sweat, saliva, and sputum<br>Urine, and feces<br>Semen, vaginal secretions, Milk | Thyroid gland<br>Heart<br>Adipose tissue<br>Gingiva<br>Prostate and testis<br>Tears, sweat, saliva, and sputum<br>Urine, and feces<br>Semen, vaginal secretions, Breast milk |

\*Applies to surgical instruments and certain invasive equipment such as endoscopes and laryngoscope blades

The second incident involved a middle-aged patient who presented with psychiatric symptoms and cognitive decline. The patient, who was an avid hunter and consumed venison and squirrel brains, also developed impaired gait. An MRI showed bilateral pulvinar signs. The EEG did not reveal periodic sharp waves, but showed nonspecific abnormalities including intermittent rhythmic delta activity. The patient required several diagnostic and therapeutic procedures including endoscopies and endotracheal intubation that entailed exposure to lymphoid tissue. Approximately 2 weeks after admission, 14-3-3, Tau protein, and RTQuIC results became available and were positive, creating a dilemma for reprocessing critical and semi-critical equipment. After several months in the hospital, the patient was discharged to a long-term care facility, and died more than 11 months after symptom onset. vCJD could not be definitively excluded until autopsy results became available several months later and revealed sCJD.

Against this backdrop, we sought to enhance our efforts to detect and prevent iatrogenic spread of TSE, including vCJD. We also aimed to increase awareness among all providers, prevent over-reaction or unnecessarily discarding expensive equipment, while simultaneously not missing an opportunity to prevent avoidable exposures.

## Methods

Rochester Regional Health System consists of 8 hospitals and 9 urgent care centers in St Lawrence County, and the 9 counties comprising the Finger Lakes region of NY. The system serves diverse populations including refugee communities and permanent or temporarily relocated citizens or expatriates from many countries. In 2018, requests for CSF analysis for suspected TSE increased 20% compared to the preceding four years.

The two-part intervention consisted of an educational campaign nearly identical to the one successfully used for device-related infections and described previously.^9^ The second component entailed and an extensive revision of our existing CJD policy, incorporating guidelines from the UK Department of Health and Social Care Advisory Committee and the World Health Organization regarding vCJD.^10, 11^ Briefly, the educational campaign included one-on-one engagements between infection preventionists (IPs), the hospital epidemiologist, the neurologist champion and frontline nurses and providers. Modes of delivery included individual face-to-face meetings, team huddles, committee meetings, group lectures and grand rounds. Relevant guidelines were posted on hospital websites and intranet.

One goal of the policy revision was early notification of all teams involved in the care including physicians, nursing staff including laboratory and sterile processing. Another goal was a simplified classification scheme of potentially infected tissue. Tissues were divided into three categories depending on their risk of infectivity: high, low, and no detectable infectivity (Table Only single use or disposable instruments are recommended for suspected cases who undergo surgery or invasive procedures involving contact with other than “no-risk” tissues. Non-disposable instruments or durable medical equipment that contact other than no risk tissues should be quarantined for possible decommissioning with appropriate labeling. All instruments are considered potentially contaminated when used on patients with conditions not clearly diagnosed before the procedure, and those instruments are either reprocessed according to prion guidelines or quarantined until the diagnosis is confirmed.

## Results

During the 2-year follow up period, there were no similar incidents or other near misses. In response to queries from other IPs and epidemiologists, the policy was shared with hospitals outside of the network. Feedback was uniformly positive.

## Discussion

TSEs present diagnostic, infection control and patient notification challenges. Most clinicians are unfamiliar with TSEs due to the rarity of those diseases. vCJD and CJD can be hard to distinguish clinically. No gold standard laboratory test exists for differentiating types of TSEs. The turn-around-time for RT-QiC can be up to 14 days or longer. In some cases, definitive diagnosis requires brain biopsy or only becomes available after autopsy. Last, the National Prion Center typically only releases autopsy results to public health departments, furthering diagnostic delay.

This approach could be criticized for not being warranted or justifiable due to the fact vCJD is extraordinarily uncommon. Even though sCJD is far more common than vCJD in the U.S., patients with suspected vCJD have been increasingly encountered in our health system. The suspicion was based on epidemiologic exposures and symptoms or signs consistent with vCJD. For example, having lived in the United Kingdom or France for several years or decades, or occupational exposure as taxidermists or abattoirs, or regular consumption of animal brains or cervid meat products.Furthermore, available data indicate that the incidence of CJD in cervids is increasing, and the potential exists for transmission to humans and subsequent human disease, and the species barrier is not static.^7^ These occurrences are unlikely to be unique to our healthcare system. To that end, we make our policy to those interested. It is available from the corresponding author upon request.

## Data Availability

NA

## Acknowledgements

None

